# Psychological Distress and Post-Traumatic Stress Symptoms among Medical Students in Vietnam: Associations with Non-Contact and Contact Sexual Violence Exposure

**DOI:** 10.64898/2026.07.25.26358649

**Authors:** Phan Thi Minh Ngoc, Hoang Thi Hai Van, Tran Quynh Anh, Leah E. Daigle, Doan Ngoc Thuy Tien, Kathryn M. Yount

## Abstract

**Background:** Medical students face significant mental health challenges, yet psychological distress and post-traumatic stress remain insufficiently characterized in lower-resource settings like Vietnam. Among potential stressors, sexual violence (SV) exposure represents a critical but understudied risk factor. This study evaluated the severity of psychological distress (PD) and post-traumatic stress symptoms (PTSS) among medical students in Northern Vietnam and examined their associations with past-year non-contact and contact SV exposure.

**Methods:** A cross-sectional survey was conducted in May 2025 using REDCap among a probability sample of 555 medical students (years 2–5). Mental health outcomes were evaluated using validated instruments: the 21-item Depression, Anxiety, and Stress Scales (DASS-21) for past-week PD and the PTSD Checklist for DSM-5 (PCL-5) for past-month PTSS. Past-year non-contact and contact SV exposures were measured using the Sexual Experiences Survey–Victimization (SES-V). Multivariable linear regression modeled adjusted associations between SV exposure types and mental health symptom severity, testing for sex-specific differences.

**Results:** Overall mental health symptom scores escalated significantly with SV exposure. Median PTSS scores increased from 11 among unexposed students to 20 for non-contact SV and 30 for contact SV (*p* < 0.001). Similarly, median PD scores increased from 7 (unexposed) to 11.5 (non-contact SV) and 21 (contact SV) (*p* < 0.001). Adjusted linear models demonstrated robust associations where both non-contact and contact SV exposure were strong independent predictors of heightened PD and PTSS severity, with no significant moderation by sex. In terms of exposure prevalence, 21.26% of students reported non-contact SV (higher in females: 26.24% vs. males: 16.12%) and 10.66% reported contact SV.

**Conclusions:** Vietnamese medical students exposed to sexual violence experience a heavy mental health burden marked by elevated psychological distress and post-traumatic stress symptoms. To protect student mental health in lower-resource settings, medical institutions must establish comprehensive trauma-informed psychological support, confidential counseling services, and targeted intervention strategies addressing both contact and non-contact stress exposures.

**Highlights:**

- Prior-year non-contact and contact sexual violence are common in medical students in Northern Vietnam.
- Non-contact sexual violence is more common among female than male students.
- Contact sexual violence is more common among sexual-minority than heterosexual students.
- Non-contact and especially contact sexual violence are positively associated with psychological distress and post-traumatic stress.
- Inclusive, preventive and response services are needed to address sexual violence and its psychological impacts.

## 1. Introduction

*Sexual violence,* broadly defined as any sexual act, attempt to obtain a sexual act, or unwanted sexual comments or advances directed against an individual’s sexuality through coercion, regardless of the setting or the relationship to the victim is a public health concern and a violation of human rights.^1^ Sexual violence victimization is generally categorized into two domains: contact and non-contact violence. Contact sexual violence involves any non-consensual physical interaction, ranging from unwanted sexual touching to attempted or completed rape involving physical force or incapacitation.^1,2^ Non-contact sexual violence includes behaviors that do not involve direct physical contact but are intrusive, such as verbal sexual harassment, exhibitionism, and other forms of unwanted sexual conduct.^3,4^ A comprehensive understanding of these forms is essential for assessing the prevalence of sexual violence among university students, a population known to be at elevated risk globally.^5^

In the United States, approximately one in five college women experience sexual violence, with 23.1% reporting incidents involving physical force or incapacitation during their studies.^2^ Similar patterns have been observed in Europe, where 36.7% of university students in France and 9.1% of female students in Sweden report recent sexual harassment.^6,7^ In Sub-Saharan Africa, the pooled lifetime prevalence of sexual violence among female youth in educational institutions is 26.2%, reaching nearly 50% in certain regions.^8,9^ Despite these high prevalences, sexual violence is considered to be substantially underreported, particularly among sexual and gender minoritized students due to stigma, fear of retaliation, and lack of trust in institutional systems.^10,11^

In Southeast Asia, the lifetime prevalence of sexual victimization among youth ranges from 3% to 65% for girls and from 3% to 42% for boys.^12^ In Indonesia, where studies of sexual violence are more common, 56.8% of university students report experiences of sexual violence, predominantly non-contact (verbal or emotional) forms.^3^ In Vietnam, sexual violence also is prevalent and is thought to be driven by longstanding norms that emphasize male sexual entitlement and perpetuate victim-blaming.^13,14^ The epidemiology of violence among Vietnamese youth is characterized by high levels of poly-victimization, with 94.3% of high school students reporting lifetime exposure to at least one form of violence,^15^ and 31.1% experiencing more than ten different forms.^16^ Moreover, a substantial reporting gap exists, while women report victimization at rates of around 12%, men’s self-reported perpetration rates are as low as 0.2%, highlighting potentially narrow definitions, tendencies to dismiss, or cultural barriers disclosing violent behavior.^13^

The consequences of sexual violence can be profound and longlasting. Sexual violence victimization is a strong and consistent predictor of severe depression and anxiety.^7^ In longitudinal studies, victims exhibit more depressive symptoms, especially after non-consensual sexual experiences.^7,17^ In Vietnam, baseline rates of depression among medical students already are high (15.2%), placing sexual violence victims at heightened risk of compounded mental-health burdens.^18^ Beyond depression, sexual violence victimization is a major precipitating factor for post-traumatic stress disorder (PTSD) and acute stress reactions.^19^ Experiences involving coercion or physical force often lead to emotional dysregulation, with victims showing elevated levels of psychotic symptoms and clinical anxiety.^19^ Unaddressed, this cumulative psychological burden can contribute to substance use and suicidal ideation and planning^20^ and can disrupt academic performance and contribute to school dropout.^21^

Although the prevalence of sexual violence is increasingly documented among university students, university surveys of sexual violence remain sparse in lower-resource settings.^22^ A critical need exists to understand the associations between different forms of sexual violence with a range of adverse mental health outcomes that coincide with and contribute to diminished academic performance at universities in Vietnam. This study aims 1) to describe and compare the prevalence of sexual violence victimization by sex and other characteristics among medical students in Northern Vietnam, and 2) to test for sex-specific associations between sexual violence victimization and symptoms of psychological distress (depression, anxiety, and stress) and post-traumatic stress.

## 2. Methods

### 2.1 Study setting

The study was conducted at a medical university in Northen, Vietnam, which newly enrolls undergraduate students across various health-related professions. There are two undergraduate tracks: a bachelor’s program lasting four years and a doctor’s program lasting six years. All programs begin with two years of on-campus study, followed by a clinical phase, during which students spend most of their time in hospitals. A substantial proportion of students come from provinces outside Hanoi. Among them, about 30% live in on-campus dormitories, while the remainder live in boarding houses near the university to reduce living costs. In addition to academics, students engage in various extracurricular activities that involve interaction with diverse groups of people. The university has established a student support center, which has been operating for the past two years and provides free services for students seeking support.

### 2.2 Study design

This mixed-methods cross-sectional study on Sexual and Mental health involved a quantitative survey of students and 10 post-survey in-depth interviews with survivors to understand their experiences of sexual violence victimization and help-seeking behaviors. Survey findings are reported here.

### 2.3 Eligibility, sample size, and sampling method

Eligible participants were all 5,000 students in years two to five at the study university who consented to participate in the online survey. For alpha=0.05 for a two-tailed test, a margin of error ε=0.05, a population size of 5,000, an estimated prevalence of sexual violence victimization of 31.5%,^13^ the required sample size was 312. Expecting ∼80% participation,^13^ and assuming a design effect of 1.2, the smallest sample size should be 312*1.2=375. To be conservative, we aimed to recruit 600 participants in the survey. To do so, we compiled a list of student identification numbers for all students from Years 2 to 5 and applied stratified random sampling to ensure that the selected sample reflected the distribution of students across academic years. Using this approach, 600 eligible students were randomly selected within strata and invited to participate.

### 2.4 Recruitment, participants, and procedure

This cross-sectional study was conducted during the 2025–2026 academic year. The Department of Student Affairs and Dormitory Management provided the email addresses of selected students. Two days before launching the survey, an initial email was sent to inform students about the study as an official, university-approved activity. Subsequently, invitation emails containing the REDCap survey link were distributed. Students accessed an online consent form, read it, and electronically signed it before entering the survey. For students who had not opened the link or had opened it but not completed the survey, REDCap automatically sent reminder emails every two days for 14 days. The university’s student management system also reminded students to check their email and participate in the study.

All 600 invited students opened the survey link; 45 (7.5%) declined participation; and 555 (92.5%) completed the survey. At the end of the questionnaire, students who reported sexual violence victimization were invited to participate in in-depth interviews to further explore their experiences and help-seeking behaviors. A total of 179 participants expressed interest, of whom 10 were selected, and 10 interviews were completed.

Upon opening the personalized REDCap link, participants were provided an information sheet explaining the study, along with the principal investigator’s phone number and email address for questions. Consenting participants received several survey modules in sequence. A short demographics module gathered information on age, sex assigned at birth, sexual orientation, ethnicity, living arrangement, and prior relationship/sexual experience. The 20-item PTSD Checklist for DSM-5 (PCL-5) then assessed post-traumatic stress symptoms (PTSS) related to intrusion (5 items), avoidance (2 items), negative alterations in cognition and mood (7 items), and alterations in arousal and reactivity (6 items).^23^ The 21-item Depression, Anxiety, and Stress Scale (DASS-21) ^24^ assessed symptoms of depression (7 items), anxiety (7 items), and stress (7 items). The Sexual Experiences Survey – Victimization (SES-V) assessed exposure to non-contact (10 items) and contact 35 items) sexual violence in the past 12 months. At the end of the survey, all participants received a resource list and asked to report their level of distress and its manageability. Those reporting the highest distress (=10) or unmanageable distress regardless of the level and who would like confidential follow-up were referred to a non-study staff-trained counselor for assessment and referral. Mean survey time among completers was 20 minutes, and mean in-depth interview time was 30 minutes. Participants in the survey received $4, and participants in the in-depth interviews received $16.

### 2.5 Mental-health outcomes

Post-traumatic stress symptoms (PTSS) were assessed using the 20-item PCL-5. Respondents reported how much they have been bothered by each symptom in the prior month, using the rating scale 0 “Not at all,” 1 “A little bit, 2 “Moderately,” 3 “Quite a bit,” and 4 “Extremely,” reflecting a change from 1-5 in the DSM-IV version. Example symptoms included “repeated, disturbing, and unwanted memories of the stressful experience?” The PCL-5 has been used to screen for PTSS symptoms in China and Vietnam.^25,26^ The internal consistency (Cronbach’s alpha) of the PCL-5 total score was 0.951 in this sample, and all corrected item-total correlations were above 0.3, specifically, item 8 was 0.370, and all other items were greater than 0.52.^25^ Internal consistency of symptom subscores ranged from 0.602 to 0.971.

To assess psychological distress using the 21-item DASS-21, respondents rated the extent to which they have experienced each symptom over the past week, using a 4-point Likert scale ranging from 0 “Did not apply to me at all,” 1 “Applied to me to some degree, or some of the time,” 2 “Applied to me to a considerable degree, or a good part of time,” and 3 “Applied to me very much, or most of the time.” Example items include “I felt down-hearted and blue” and “I found it hard to relax.” Scores on three subscales naming DASS-21-Depression (DASS-21-D), DASS21-Anxiety (DASS-21-A) and Stress (DASS-21-S) can then be calculated. The DASS-21 is reliable and suitable for use to assess symptoms of common mental health problems, especially depression and anxiety among Vietnamese adolescents.^27^ In this sample, internal consistency was 0.91 for the DASS-21 total score and 0.82 – 0.94 for the subscores.

### 2.6 Primary exposure

The primary exposure was sexual violence experience in past 12 months measured using The Sexual Experience Survey Victimization (SES-V). The Sexual Experiences Survey – Victimization (SES-V, 2024) is a 45-item self-report measure that assesses experiences of sexual violence victimization across two domains: non-contact sexual violence (10 items) and contact sexual violence (35 items). The non-contact domain includes behaviors such as unwanted sexual comments, exposure, and coerced sexual acts without physical contact, while the contact domain covers a range of unwanted physical experiences, from sexual touching to attempted or completed penetration, using tactics such as verbal pressure, manipulation, or physical force. Respondents report the frequency of each experience within the past 12 months using response options of 0 “Never,” 1 “Once,” 2 “Twice,” and 3 “Three or more times.” Example items included “Someone made sexual comments or gestures toward me without my consent” and “Someone fondled, kissed, or touched me in a sexual way without my consent”.^28^ SES Vietnamese version was validated and has been applied to Vietnamese male students.^13^

### 2.7 Covariates

The analysis controlled for the following covariates:^13^ sexual orientation (heterosexual, sexual minority),^29^ age (age was categorized into four quartiles (Q1-Q4) based on the sample distribution to protect participant anonymity), living arrangement (with parents/relatives, on campus, off campus alone or with non-relatives, other), ethnicity (Kinh, other ethnic minority), prior relationship (never, ever), and prior sexual activity by sex of partner (male only, female only, male and female, never).

### 2.8 Statistical analysis

Descriptive statistics were used to summarize participants’ characteristics and study variables. Categorical variables were presented as frequency distributions and percentages, and differences across groups were assessed using Pearson’s chi-square test or Fisher’s exact test, as appropriate.

The PCL-5 and DASS-21 scores and subscores were right-skewed, so these variables were summarized using medians and interquartile ranges (IQRs), and group differences were examined using the Kruskal–Wallis test. Post hoc pairwise comparisons were conducted using the Wilcoxon rank-sum tests with Bonferroni correction to account for multiple comparisons. Given that three pairwise comparisons were performed (i.e. no sexual violence vs. non-contact sexual violence, no sexual violence vs. contact sexual violence, and non-contact vs. contact sexual violence), the significance level was adjusted to α = 0.017 (0.05/3).

To examine associations between prior-12-month sexual violence exposure and mental health outcomes, multivariable OLS regression models were conducted with four models. Model 1 included sexual violence exposure and covariates as predictors of the DASS-21 score and all subscores, and Model 2 included interaction terms between sexual violence exposure and sex assigned at birth to assess potential effect modification. In turn, Model 3 included sexual violence exposure and covariates as predictors of the the PCL-5 score and all subscores, and Model 4 included interaction terms between sexual violence exposure and sex assigned at birth to assess potential effect modification. All models controlled for age, ethnicity, sexual orientation, prior relationship, living arrangement, and sexual partner history. A *p*-value<0.05 was considered statistically significant in the regression models. All analyses were conducted using Stata 14.0.

### 2.9 Ethical considerations

The University Institutional Review Board approved the study (1958/GCNHMUIRB). All identifiable information was stored separately from survey responses, and user-defined access to identifiers was restricted to non-study staff who did not have access to the survey responses. The PI conducted in-depth interviews in a trauma-informed manner to minimize any emotional distress. Participants in the in-depth interviews were assigned pseudonyms, and all identifying details were removed from the transcripts. Data were stored securely in password-protected files and were only accessible to the research team. After transcription, the audio file was held securely, accessible to non-study staff. All participants were provided with a list of mental health support services available to them.

## 3. Results

Table 1 provides the characteristics of participants, overall and by sex assigned at birth. Of 555 participants, 273 (49.19%) were male, and 282 (50.81%) were female. Participants were relatively evenly distributed across the four age quartiles, with the largest proportion falling into the first quartile (33.51%). Additionally, most students were heterosexual (90.45%). Most often, students were of Kinh ethnicity (88.11%), had ever been in a relationship (52.79%), and were living off campus alone or with non-relatives (44.86%). Male and female students differed in age, prior relationship, and prior sexual experience (Table 1), with male students more often being older, having ever had a relationship, and having ever had sex (Table 1).

**Table 1.** Demographic characteristics of participants, 555 second to fifth year students at a Medical University in Northern Vietnam.

| Characteristic |  | Overall<br>n (%) | Sex assigned at birth |  | p-value |
| --- | --- | --- | --- | --- | --- |
|  |  |  | Male<br>n (%) | Female<br>n (%) |  |
|  |  | n = 555 | n = 273 | n = 282 |  |
| **Age (Quartiles)** |  |  |  |  | 0.021 |
|  | Q1 | 186 (33.51) | 80 (29.30) | 106 (37.59) |  |
|  | Q2 | 145 (26.13) | 65 (23.81) | 80 (28.37) |  |
|  | Q3 | 102 (18.38) | 57 (20.88) | 45 (15.96) |  |
|  | Q4 | 122 (21.98) | 71 (26.01) | 51 (18.09) |  |
| **Sexual orientation** |  |  |  |  | 0.577 |
|  | Heterosexual | 502 (90.45) | 245 (89.74) | 257 (91.13) |  |
|  | Sexual minority | 53 (9.55) | 28 (10.26) | 25 (8.87) |  |
| **Ethnicity** |  |  |  |  | 0.903 |
|  | Kinh | 489 (88.11) | 241 (88.28) | 248 (87.94) |  |
|  | Non-Kinh | 66 (11.89) | 32 (11.72) | 34 (12.06) |  |
| **Prior relationship/sexual activity** |  |  |  |  | 0.001 |
|  | Never | 262 (47.21) | 109 (39.93) | 153 (54.26) |  |
|  | Ever | 293 (52.79) | 164 (60.07) | 129 (45.74) |  |
| **Living arrangement** |  |  |  |  | 0.914 |
|  | With parents/relatives | 146 (26.31) | 74 (27.11) | 72 (25.53) |  |
|  | On-campus | 152 (27.39) | 75 (27.47) | 77 (27.30) |  |
|  | Off campus alone or<br>with non-relatives | 249 (44.86) | 121 (44.32) | 128 (45.39) |  |
|  | Others | 8 (1.44) | 3 (1.10) | 5 (1.77) |  |
| **Have you ever had sex with** |  |  |  |  | **<0.0001** |
|  | Male only | 61 (10.99) | 14 (5.13) | 47 (16.67) |  |
|  | Female only | 66 (11.89) | 62 (22.71) | 4 (1.42) |  |
|  | Male and female | 9 (1.62) | 5 (1.83) | 4 (1.42) |  |
|  | Never | 419 (75.50) | 192 (70.33) | 227 (80.50) |  |

*Note: P-values were derived from Pearson's chi-square test or Fisher's exact test. Figures in bold indicate Fisher's exact texts. Source: Own calculations.*

Table 2 presents the distribution of sexual violence exposure in the prior 12 months with categories of covariates. The prevalence of prior-year sexual violence differed by sex, with no exposure being more common among males (72.89%) than females (63.48%), non-contact sexual violence being more common among females (26.24%) than males (16.12%), and contact sexual violence being similarly prevalent between females (10.28%) and males (10.99%). Otherwise, most notably, the prevalence of prior-year sexual violence differed significantly by sexual orientation, prior relationship experience, and prior sexual activity. Any contact sexual violence was more common among sexual minority students (50.94%) than heterosexual students (6.37%), and no prior-year sexual violence was much more common among heterosexual students (71.91%) than sexual minority students (32.08%). Contact sexual violence was more common for students who had been in a relationship versus not (16.38% versus 4.20%), who had had prior sexual activity versus not (16.38% versus 4.2%), and who had had sex previously with a male partner (27.87% male partner only; 77.78% male and female partner only).

**Table 2:** Distribution of Exposure to Sexual Violence in the Prior 12 Months with Covariate Categories, 555 2^nd^-5^th^ Year Undergraduate Students Attending a Medical University in Northern Vietnam.

| Characteristic | No Sexual Violence<br>n (%) | Non-contact Sexual Violence only<br>n (%) | Any Contact Sexual Violence<br>n (%) | p-value |
| --- | --- | --- | --- | --- |
|  | **n = 378** | **n=118** | **n= 59** |  |
| **Sex assigned at birth** |  |  |  | 0.014 |
| Male | 199<sup>a</sup> (72.89) | 44<sup>a</sup> (16.12) | 30<sup>a</sup> (10.99) |  |
| Female | 179<sup>b</sup> (63.48) | 74<sup>b</sup> (26.24) | 29<sup>b</sup> (10.28) |  |
| **Age (Quartiles)** |  |  |  | 0.091 |
| Q1 | 128<sup>a</sup> (68.82) | 42<sup>a</sup> (22.58) | 16<sup>a</sup> (8.60) |  |
| Q2 | 96<sup>a</sup> (66.21) | 29<sup>a</sup> (20.00) | 20<sup>a</sup> (13.79) |  |
| Q3 | 80<sup>a</sup> (78.43) | 16<sup>a</sup> (15.69) | 6<sup>a</sup> (5.88) |  |
| Q4 | 74<sup>a</sup> (60.66) | 31<sup>a</sup> (25.41) | 17<sup>a</sup> (13.93) |  |
| **Sexual orientation** |  |  |  | **<0.0001** |
| Heterosexual | 361<sup>a</sup> (71.91) | 109<sup>a</sup> (21.71) | 32<sup>a</sup> (6.37) |  |
| Sexual minority | 17<sup>b</sup> (32.08) | 9<sup>b</sup> (16.98) | 27<sup>b</sup> (50.94) |  |
| **Ethnicity** |  |  |  | 0.235 |
| Kinh | 327<sup>a</sup> (66.87) | 108<sup>a</sup> (22.09) | 54<sup>a</sup> (11.04) |  |
| Non-Kinh | 51<sup>b</sup> (77.27) | 10<sup>b</sup> (15.15) | 5<sup>b</sup> (7.58) |  |
| **Prior relationship/sexual activity** |  |  |  | **<0.0001** |
| Never | 193<sup>a</sup> (73.66) | 58<sup>a</sup> (22.14) | 11<sup>a</sup> (4.20) |  |
| Ever | 185<sup>b</sup> (63.14) | 60<sup>b</sup> (20.48) | 48<sup>b</sup> (16.38) |  |
| **Living arrangement** |  |  |  | **0.659** |
| With parents/relatives | 98<sup>a</sup> (67.12) | 34<sup>a</sup> (23.29) | 14<sup>a</sup> (9.59) |  |
| On campus | 100<sup>a</sup> (65.79) | 38<sup>a</sup> (25.00) | 14<sup>a</sup> (9.21) |  |
| Off campus alone or with non-relatives | 175<sup>a</sup> (70.28) | 44<sup>a</sup> (17.67) | 30<sup>a</sup> (12.05) |  |
| Others | 5<sup>b</sup> (62.50) | 2<sup>b</sup> (25.00) | 1<sup>b</sup> (12.50) |  |
| **Ever had sex with** |  |  |  | **<0.0001** |
| Male only | 28<sup>a</sup> (45.90) | 16<sup>a</sup> (26.23) | 17<sup>a</sup> (27.87) |  |
| Female only | 47<sup>a</sup> (71.21) | 10<sup>a</sup> (15.15) | 9<sup>a</sup> (13.64) |  |
| Male and female | 2<sup>b</sup> (22.22) | 0<sup>b</sup> (0.00) | 7<sup>b</sup> (77.78) |  |
| Never | 301<sup>b</sup> (71.84) | 92<sup>b</sup> (21.96) | 26<sup>b</sup> (6.21) |  |

Note: P-values were derived from Pearson's chi-square test or Fisher's exact test. Figures in bold indicate Fisher's exact test. Post hoc pairwise comparisons were conducted using the Bonferroni correction for multiple comparisons ( $\alpha = 0.017$ ). Superscript letters (a, b) indicate results of pairwise comparisons, values sharing the same letter are not significantly different. Source: Own calculations.

Table 3 presents the distribution of the PCL-5 score, DASS-21 score, and their subscale scores among categories of sexual violence exposure in the prior 12 months. For post-traumatic stress symptoms (PTSS), the median overall PCL-5 score was 11 (IQR: 3–22) among students with no sexual violence, 20 (IQR: 8–36) among those with non-contact sexual violence only, and 30 (IQR: 20–40) among those with contact sexual violence. A similar pattern was observed across all PTSD subscales, including intrusion, avoidance, negative alterations in cognition and mood, and alterations in arousal and reactivity. For psychological distress, the median DASS-21 scores were 7 (IQR: 2–17), 11.5 (IQR: 6.5–20), and 21 (IQR: 15–30) across the no sexual violence, non-contact sexual violence only, and any-contact sexual violence groups, respectively. For all scores and subscores of psychological distress and post-traumatic stress symptoms, there was a clear and significant dose-response association with prior-year sexual violence, with unexposed students having the lowest scores, those exposed to non-contact sexual violence only having intermediate scores, and those exposed to contact sexual violence having the highest scores. Post hoc pairwise comparisons with Bonferroni correction indicated that mental health conditions were significantly different among the three sexual violence exposure groups, as denoted by differing superscript letters.

**Table 3:**
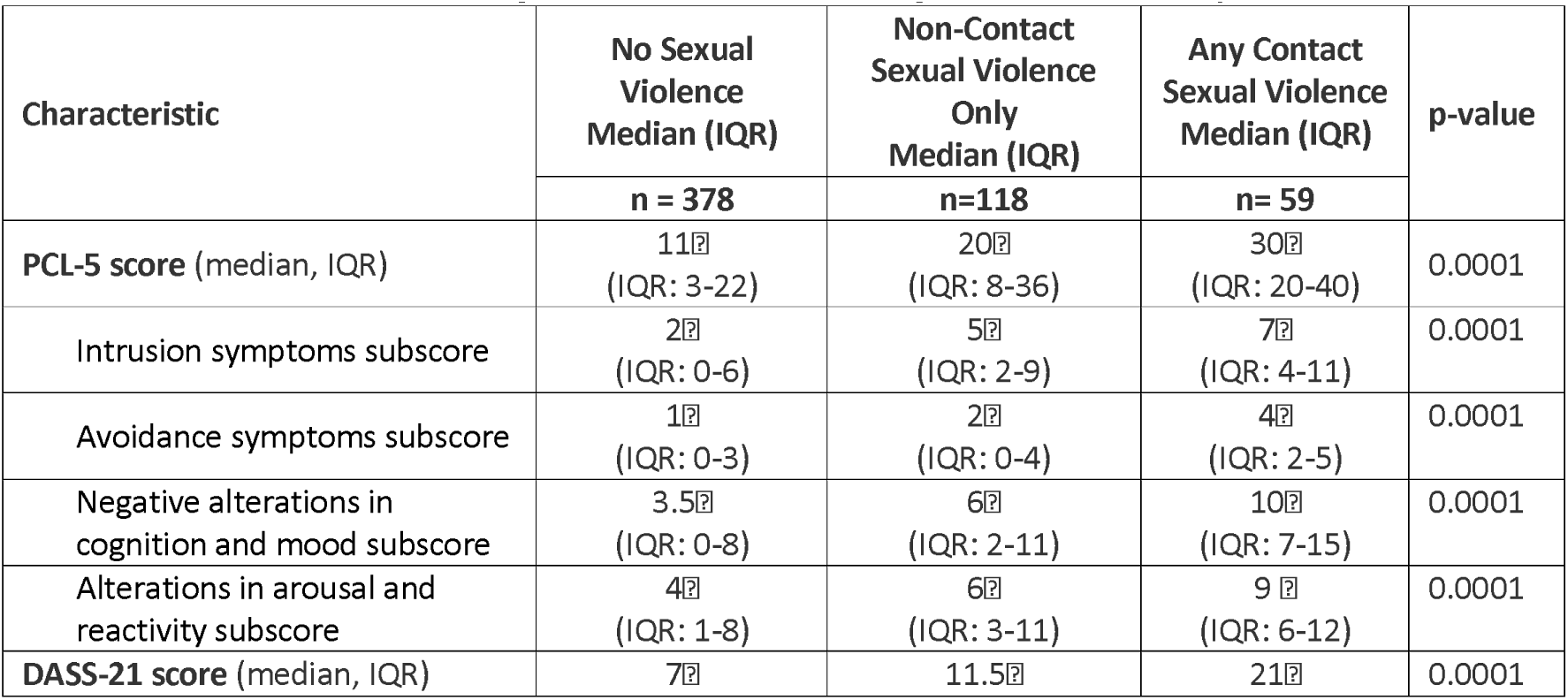
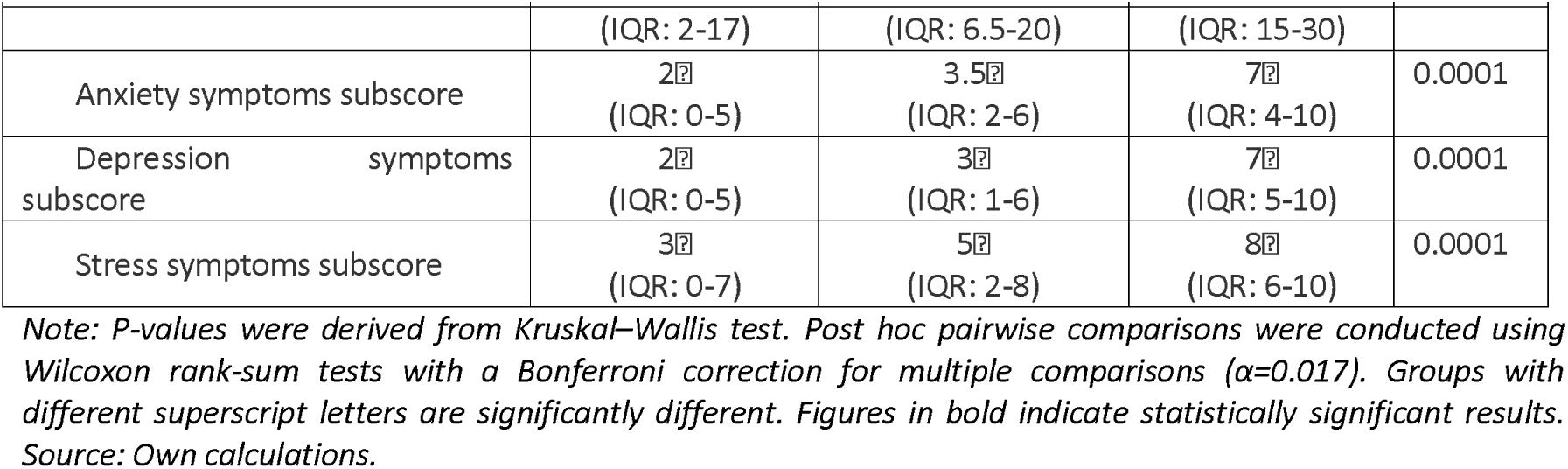
Distribution of mental health conditions by Category of Exposure to Sexual Violence in the Prior 12 Months, 555 2^nd^-5^th^ Year Undergraduate Students Attending a Medical University in Northern Vietnam.

Table 4 presents the adjusted associations of sexual violence with the DASS-21 overall score and subscale scoresfor anxiety, depression, and stress. In Model 1 (without interaction terms), exposure to sexual violence was consistently associated with higher levels of psychological distress, including anxiety, depression, and stress. Compared with students reporting no sexual violence, those experiencing non-contact sexual violence only in the prior year had significantly higher anxiety (β=1.197, 95% CI: 0.536–1.859), stress (β=1.417, 95% CI: 0.601–2.233), and overall DASS-21 (β=3.228, 95% CI: 1.248–5.209) scores. The association between non-contact sexual violence and depression was not statistically significant.

**Table 4.** Multivariable Logistic Regressions for the Association of Non-Contact Sexual Violence and Contact Sexual Violence with Subscores for Symptoms of Anxiety, Depression, and Stress, and with the Overall DASS-21 Among Students at Medical University in North Vietnam

| Characteristics | Anxiety score |  | Depression score |  | Stress score |  | Overall DASS-21 score |  |
| --- | --- | --- | --- | --- | --- | --- | --- | --- |
|  | Coef | 95%CI | Coef | 95%CI | Coef | 95%CI | Coef | 95%CI |
| **Model 1 (without interaction terms)** |  |  |  |  |  |  |  |  |
| **Sexual violence** (ref: none) |  |  |  |  |  |  |  |  |
| Non-contact sexual violence only | **1.197\*\*\*** | 0.536, 1.859 | 0.613 | -0.083, 1.310 | **1.417\*\*** | 0.601, 2.233 | **3.228\*\*** | 1.248, 5.209 |
| Any contact sexual violence | **4.318\*\*\*** | 2.932, 5.705 | **3.913\*\*\*** | 2.442, 5.384 | **3.474\*\*\*** | 1.989, 4.960 | **11.707\*\*\*** | 7.596, 15.818 |
| **Gender** (ref: Male) |  |  |  |  |  |  |  |  |
| Female | 0.403 | 0-0.164, 0.972 | 0.144 | -0.478, 0.767 | 0.561 | -0.126, 1.248 | 1.109 | -0.633, 3.410 |
| **Model 2 (with interaction terms)** |  |  |  |  |  |  |  |  |
| **Sexual violence** (ref: none) |  |  |  |  |  |  |  |  |
| Non-contact sexual violence only | **1.419\*\*** | 0.498, 2.341 | 0.898 | -0.155, 1.953 | **1.807\*\*** | 0.591, 3.023 | **4.127\*\*** | 1.219, 7.035 |
| Any contact sexual violence | **4.764\*\*\*** | 2.664, 6.865 | **4.570\*\*\*** | 2.337, 6.802 | **4.060\*\*\*** | 1.930, 6.190 | **13.396\*\*\*** | 7.190, 19.601 |
| **Sex assigned at birth** (ref: Male) |  |  |  |  |  |  |  |  |
| Female | 0.583 | -0.089, 1.256 | 0.393 | -0.357, 1.144 | **0.830\*** | 0.003, 1.657 | 1.809 | -0.286, 3.906 |
| **Sexual violence \* Sex assigned at birth** |  |  |  |  |  |  |  |  |
| Non-contact sexual violence only * Female | -0.397 | -1.689, 0.894 | -0.514 | -1.914, 0.886 | -0.687 | -2.324, 0.949 | -1.601 | -5.538, 2.336 |
| Any contact sexual violence * Female | -0.892 | -3.272, 1.488 | -1.216 | -3.701, 1.076 | -1.176 | -3.525, 1.172 | -3.383 | -10.064, 2.336 |

*Note: OLS regression results, controlling for demographic variables (including age, ethnicity, have you ever had sex with, living arrangement, sexual orientation). Figures in bold indicate statistically significant results. \* $p < 0.05$ , \*\* $p < 0.01$ , \*\*\* $p < 0.001$ . Source: Own calculations.*

Experiencing any contact sexual violence in the prior year was significantly associated with all mental health outcomes. Specifically, those exposed to contact sexual violence had significantly higher anxiety (β=4.318, 95% CI: 2.932–5.705), depression (β=3.913, 95% CI: 2.442–5.384), stress (β=3.474, 95% CI: 1.989–4.960), and overall DASS-21 scores (β=11.707, 95% CI: 7.596–15.818). Gender was not associated with these outcomes in Model 1.

In Model 2 (with interaction terms), the association between sexual violence and mental health conditions remained unchanged (Table 4). Except for depression, non-contact sexual violence remained significantly associated with higher anxiety (β=1.419, 95% CI: 0.498–2.341), stress (β=1.807, 95% CI: 0.591–3.023), and overall DASS-21 scores (β=4.127, 95% CI: 1.219–7.035). Similarly, contact sexual violence remained strongly associated with all mental health outcomes, with effect sizes slightly larger than those observed in Model 1.

Regarding sex assigned at birth, females had a higher stress score compared to males (β=0.830, 95% CI: 0.003–1.657), while no statistically significant differences in sex assigned at birth were observed for anxiety, depression, and overall DASS-21 scores. Importantly, no significant interaction effects were found between sexual violence and sex assigned at birth among any of the outcomes, suggesting that the associations between sexual violence and mental health outcomes did not differ significantly by sex.

Table 5 indicates the associations between sexual violence and PTSD-related symptoms (PTSS), including intrusion, avoidance, negative alterations in cognition and mood, alterations in arousal and reactivity, and the overall PCL-5 score. In Model 3 (without interaction terms), both non-contact and contact sexual violence were significantly associated with higher levels of PTSS across all domains. Compared with students reporting no sexual violence, those exposed to non-contact sexual violence only had significantly higher intrusion (β=1.813, 95% CI: 0.924–2.703), avoidance (β=0.701, 95% CI: 0.274–1.127), negative alterations in cognition and mood (β=1.831, 95% CI: 0.684–2.979), and alterations in arousal and reactivity (β=1.852, 95% CI: 0.899–2.806) scores. A similar pattern was observed for the overall PCL-5 score (β=6.199, 95% CI: 3.129–9.269). Exposure to contact sexual violence demonstrated substantially larger effect sizes among all PTSS symptoms. Sex assigned at birth was not significantly associated with any PTSS outcomes in Model 3.

**Table 5:** Multivariable Logistic Regressions for the Association of Non-Contact Sexual Violence and Contact Sexual Violence with Subscores for Symptoms of Intrusion, Avoidance, Negative Alterations, Alterations, and with the Overall PCL-5 Among Students at Medical University in North Vietnam

| Characteristics | Intrusion symptoms |  | Avoidance symptoms |  | Negative alterations in cognition and mood |  | Alterations in arousal and reactivity |  | Overall PCL-5 score |  |
| --- | --- | --- | --- | --- | --- | --- | --- | --- | --- | --- |
|  | Coef | 95%CI | Coef | 95%CI | Coef | 95%CI | Coef | 95%CI | Coef | 95%CI |
| **Model 3 (without interaction terms)** |  |  |  |  |  |  |  |  |  |  |
| **Sexual violence (ref: None)** |  |  |  |  |  |  |  |  |  |  |
| Non-contact only | **1.813\*\*\*** | 0.924, 2.703 | **0.701\*\*\*** | 0.274, 1.127 | **1.831\*\*** | 0.684, 2.979 | **1.852\*\*\*** | 0.899, 2.806 | **6.199\*\*\*** | 3.129, 9.269 |
| Any contact | **4.149\*\*\*** | 2.514, 5.785 | **2.673\*\*\*** | 1.190, 2.366 | **5.607\*\*\*** | 3.576, 7.639 | **4.135\*\*\*** | 2.407, 5.864 | **15.671\*\*\*** | 10.039, 21.304 |
| **Sex assigned at birth (ref: Male)** |  |  |  |  |  |  |  |  |  |  |
| Female | -0.068 | -0.779, 0.641 | 0.087 | -0.231, 0.405 | -0.346 | -1.278, 0.586 | 0.121 | -0.676, 0.919 | -0.206 | -2.710, 2.298 |
| **Model 4 (with interaction terms)** |  |  |  |  |  |  |  |  |  |  |
| **Sexual violence (ref: none)** |  |  |  |  |  |  |  |  |  |  |
| Non-contact only | **2.187\*\*** | 0.767, 3.606 | **0.741\*\*** | 0.127, 1.355 | 1.792 | -0.030, 3.615 | **2.024\*\*** | 0.507, 3.542 | **6.746\*\*** | 1.968, 11.524 |
| Any contact | **5.140\*\*\*** | 2.813, 7.468 | **2.137\*\*\*** | 1.277, 2.998 | **6.244\*\*\*** | 3.253, 9.236 | **4.925\*\*\*** | 2.391, 7.460 | **18.449\*\*\*** | 10.153, 26.744 |
| **Sex assigned at birth (ref: Male)** |  |  |  |  |  |  |  |  |  |  |
| Female | 0.286 | -0.526, 1.099 | 0.183 | -0.197, 0.564 | -0.213 | -1.315, 0.888 | 0.361 | -0.591, 1.315 | 0.618 | -2.351, 3.589 |
| **Sexual violence \* Sex assigned at birth** |  |  |  |  |  |  |  |  |  |  |
| Non-contact sexual violence only * Female | -0.677 | -2.507, 1.153 | -0.086 | -0.934, 0.761 | 0.033 | -2.311, 2.378 | -0.328 | -2.296, 1.638 | -1.058 | -7.316, 5.200) |
| Any contact sexual violence * Female | -1.983 | -4.530, 0.562 | -0.715 | -1.709, 0.278 | -1.261 | -4.620, 2.097 | -1.575 | -4.335, 1.184 | -5.536 | -14.515, 3.443 |

*Note: OLS regression results, controlling for demographic variables (including age, year in course, ethnicity, have you ever had sex with, living arrangement, sexual orientation). Figures in bold indicate statistically significant results. \*p<0.05, \*\*p<0.01, \*\*\*p<0.001. Source: Own calculations.*

In Model 4 (with interaction terms), the associations were mostly consistent with those in Model 3. Specifically, non-contact sexual violence continued to be significantly associated with higher intrusion (β=2.187, 95% CI: 0.767–3.606), avoidance (β=0.741, 95% CI: 0.127–1.355), alterations in arousal and reactivity (β=2.024, 95% CI: 0.507–3.542), and overall PCL-5 scores (β=6.746, 95% CI: 1.968–11.524). Similarly, contact sexual violence remained strongly associated with all PTSD symptom domains, with larger effect sizes observed for intrusion (β=5.140, 95% CI: 2.813–7.468), avoidance (β=2.137, 95% CI: 1.277– 2.998), negative alterations in cognition and mood (β=6.244, 95% CI: 3.253–9.236), alterations in arousal and reactivity (β=4.925, 95% CI: 2.391–7.460), and overall PCL-5 scores (β=18.449, 95% CI: 10.153–26.744).

No significant interaction effects were observed between sexual violence and sex assigned at birth among any PTSS, indicating that the associations between sexual violence and PTSS did not differ significantly by sex.

## 4. Discussion

Sexual violence victimization was common among medical students, with 21.26% reporting non-contact sexual violence only and 10.66% reporting contact sexual violence. However, the prevalence of non-contact sexual violence in our sample was lower than that reported in other settings, such as among female students in Eswatini, where up to 90% experienced sexualized street harassment.^30^ Similarly, a study of college students in Indonesia identified verbal abuse affecting 40.6% of the student cohort.^30^ Conversely, the 10.66% prevalence of contact sexual violence in our sample is consistent with, or slightly lower than, global estimates. In France, 36.7% of students (95% CI [31.3, 42.4]) reported experiencing at least one form of gender-based sexual violence during the past three months.^6^ In the United States, the Campus Sexual Assault Study found that 13.7% of undergraduate women reported completed sexual assault, including 4.7% involving physical force.^2^ Considerably higher rates have been reported in developing regions, such as in Nigeria where 52.5% of female students experienced unwanted touching and 19.1% reported rape.^31^ Furthermore, a comprehensive meta-analysis of Sub-Saharan African educational institutions indicated a pooled lifetime sexual violence prevalence of 26.22% among female youths.^8^

Regarding sex disparities, non-contact sexual violence was reported more often by females than males, while contact sexual violence was similar across sexes. This pattern aligns with global evidence showing that female students experience higher rates of sexual harassment.^32^ However, our finding that male and female students report similar rates of contact sexual violence adds nuance to the literature, suggesting that males are also at risk despite higher reported risks among females. This finding does support other findings that show that males are also at risk for contact sexual violence. For example, the study in Indonesia found that males reported experiencing higher rates of physical abuse (30.8%) than their female counterparts (17.8%).^3^

The high prevalence of sexual violence and sex-based differences in reporting patterns may reflect socio-cultural factors. Non-contact sexual violence toward females is often rooted in patriarchal norms, where sexualized harassment reflects and reinforces gendered power hierarchies and maintains male dominance in public and academic spaces.^17,33^ Morever, under-reporting and social desirability bias likely distort sexual violence prevalence, as only about 11% of rapes are reported to college authorities in the US.^14^ Male victimization may be underreported due to masculine norms, obscuring the true prevalence of contact sexual violence among males.^33^ Finally, the transition to university may increase sexual violence risk, as reduced supervision, academic stress, and campus social environments heighten vulnerability to both contact and non-contact sexual violence.^14,31^

Our finding that the contact sexual violence group did not differ significantly by sex adds critical nuance to the existing literature. While numerous national surveys indicate that females face a disproportionate risk for many forms of interpersonal violence,^32^ several university-based cohort studies suggest that males and females experience similar rates of physical violence and sexual coercion on campus.^19,22,34^ This suggests that while females bear more non-contact harassment, campus dynamics (e.g., hookup culture, alcohol use) may expose both sexes to similar risks of contact sexual violence.^19,32^

Other factors were related to experiencing contact sexual violence. The higher proportion of sexual minority students in the contact sexual violence group aligns with global evidence of increased vulnerability, with higher victimization rates reported among sexual minority youth than heterosexual peers (e.g., 25.3% vs. 7.6%).^11,32^ LGBTQ+ students may face intersecting homophobia, stigma, and marginalization, increasing their risk of targeted sexual violence.^10^

Moreover, students in the contact sexual violence group more often reported prior relationships and having sex with both sexes, consistent with research linking this to higher sexual violence risk.^9,30,34,35^ Having multiple recent partners are associated with higher risk of forced sex and contact sexual violence.^34,36^ Our finding that students who had sex with both sexes experienced the highest proportion of contact sexual violence (77.78%) is consistent with evidence that this group faces higher sexual violence risk than those with opposite- or same-sex partners only.^32^ These vulnerabilities likely stems from two intersecting mechanisms. First, persons with diverse sexual partner may face compounded stigma and marginalization, increasing their risk of targeted sexual violence.^10^ Second, having diverse or multiple sexual partners and previous relationships inherently increases an individual’s probability of encountering potential perpetrators.^34^

Regarding living arrangement, students living off campus alone or with non-relatives reported higher levels of contact sexual violence than those living on campus or with parents/relatives. This reality was highlighted in an interview with by a male student living off campus. He shared: *“I rent a place in a narrow alley because it’s cheaper. The way home is very dark and quiet. I was stopped, pushed against the wall, and forced to kiss and touched.”* A study among female university students in the United States suggests that contact sexual violence remains a persistent risk on campus.^37^ In contrast, a study in Nigeria found no significant difference in sexual violence prevalence between on- and off-campus students.^38^ Regardless of living arrangement, shared student cultures (e.g., partying, alcohol use, peer pressure) may increase vulnerability. On-campus systems at our selected university may offer some protection, but findings were not statistically significant and should be interpreted cautiously, requiring further research.

Our findings demonstrate a dose–response pattern in mental health (PCL-5, DASS-21), with symptoms increasing from no sexual violence to non-contact and highest in contact sexual violence, consistent with a trauma continuum.^6,21^ Contact sexual violence showed the strongest associations across all outcomes, while non-contact sexual violence was linked to anxiety, stress, and post-traumatic stress symptoms (PTSS), but not depression. This pattern is consistent with the trauma continuum, where more severe sexual violence has greater and more persistent effects. A Swedish cohort study found that, although all forms of sexual violence affected mental health, contact sexual violence (“sex against one’s will”) showed the strongest and most persistent links with severe depression and anxiety.^7^ Similarly, a prospective study of French students found that those exposed to severe or multiple forms of violence had higher depression and anxiety scores than those with less severe or isolated experiences.^6^ However, our finding that non-contact sexual violence was not significantly associated with depression contrasts with several international studies.^17,39^ The lack of association between non-contact sexual violence and depression may suggest that, although harassment increases stress and anxiety, medical students may have coping or resilience factors that buffer against depression unless exposed to contact sexual violence.^7,33^

A key finding is that both non-contact and contact sexual violence were associated with higher post-traumatic stress symptoms (PTSS) across domains (intrusion, avoidance, negative alterations in cognition/mood, and arousal/reactivity), with stronger effects for contact sexual violence. This supports evidence that sexual violence can have substantial impacts on psychological functioning, even in the absence of physical contact. This corroborates global epidemiological models indicating that sexual violence acts as a profound traumatic stressor capable of fundamentally altering psychological functioning regardless of physical contact.^37^ Non-contact sexual violence, including severe verbal coercion or intimidation, may induce emotional distress and fear sufficient to trigger post-traumatic stress responses.^19^ Data from 25 countries show that both physical and non-physical sexual violence are associated with higher PTSS and sleep problems among university students..^34^

Our analysis revealed no significant interaction between sexual violence and sex assigned at birth regarding mental health or PTSS outcomes, indicating that the psychological effects of sexual violence are similar across sex. While females reported slightly higher overall stress scores - a finding consistent with the broader literature showing that females report higher baseline internalizing symptoms^34^ - the association of sexual violence trauma on psychological health did not differ by sex. One male participant shared how his experience negatively impacted him. He noted: *“I’ve never dared to tell anyone, but I feel like everyone somehow knows what happened to me, even though that night it was only me and him. I don’t think anyone would believe me anyway—how could a guy be touched like that? I don’t want to see anyone anymore, and I feel scared all the time.”*

This pattern is consistent with a study in South-Western Uganda, where sexual coercion was similarly associated with higher anxiety, depression, and psychoticism in both males and females.^19^ Similarly, US Youth Risk Behavior Survey data show that both male and female students who experience sexual violence report comparable increases in sadness, hopelessness, and suicidality.^20^ Our findings, supported by emerging global data, confirm that when male students are victimized, they experience similarly debilitating mental health symptoms—such as PTSS, depression, and anxiety—as their female counterparts.

Ultimately, our findings corroborate the global reality that sexual violence is a pervasive issue in higher education. The relatively high rates of contact sexual violence across both sexes in the medical student population underscore the urgent need for universities to implement comprehensive, sex-inclusive prevention strategies and accessible mental health support systems on campuses.

### Limitations and Strengths

This study has some limitations. sexual violence was self-reported over 12 months and may be affected by recall and sensitivity, particularly for non-contact sexual violence. We used an anonymous online survey and behaviorally specific items to reduce this possibility. The single-university sample may limit generalizability, and small subgroups led to imprecise estimates. Findings should be interpreted cautiously and confirmed in larger studies.

Despite these limitations, this study has many strengths. Our participation rate of 92.5% (555 of 600) is higher than reported in previous studies.^17,34,38,39^ High participation may be driven by automated reminders from the REDCap system, additional outreach, small compensation, and access to support services if needed. It may also reflect the growing relevance of sexual violence as a topic among students.

In addition, this study used well-established instruments (PCL-5, DASS-21, and SES-V) that have been shown to be reliable in Vietnamese contexts. Finally, the analysis accounted for a range of relevant covariates and included interaction terms, allowing for a more robust assessment of the associations between sexual violence and mental health across sexes.

### Implications for Research and Practice

This study has several implications for research and practice. First, there is a need to recognize risk across sexes: Contact sexual violence appears similar across sexes, suggesting prevention and response efforts should be inclusive rather than gender-restricted. Second, there is a need to prioritize high-risk groups: Sexual minority students and those with more diverse partner histories may face higher risk and should be considered in targeted efforts. Third, there is a need to address the trauma continuum: The gradient in outcomes suggests both non-contact and contact sexual violence need attention—non-contact as a chronic stressor and contact sexual violence as more severe trauma. Further, there is a need to strengthen institutional responses. Universities should ensure accessible, trauma-informed services that address all forms of sexual violence and support vulnerable students.

## 5. Conclusion

Vietnamese medical students face a significant mental health burden, with psychological distress and post-traumatic stress symptoms demonstrating a clear dose–response relationship with sexual violence (SV) exposure. While non-contact SV functions as a persistent stressor associated with elevated anxiety, stress, and PTSS, contact SV correlates with more severe psychological impairment across all mental health domains, including depression. Both forms of SV consistently predict heightened PTSS regardless of sex assigned at birth. Although SV victimization is common overall, distinct vulnerability patterns emerge: females experience higher non-contact SV, while contact SV is similarly prevalent across sexes and notably higher among sexual minority students and those with more diverse sexual partner histories. Medical universities must prioritize comprehensive, trauma-informed mental health support and prevention services that address the psychological toll of all forms of sexual violence, with targeted outreach for higher-risk student populations.

## Data Availability

All data produced in the present study are available upon reasonable request to the authors

## Acknowledgements

The authors thank the participants, without whom this project would not have been possible. We also thank Tran Kim Thanh, Pham Tung Son, Tran Thi Ngoc Trang, Dao Thi Ngoan, Bui Hong Hanh, Nguyen Thi Ngoc Linh, Tran Thi Phuong, Pham Thi Thanh Huong, Tran Thi Phuong Nga for data collection support. The authors acknowledge using ChatGPT (version 5) for English grammar correction and language improvement. All suggestions were reviewed and edited by the authors, who take full responsibility for the final manuscript.

## Author CREDIT Statement

PTMN: Conceptualization, Methodology, Investigation, Data curation, Writing - Original draft

HTHV: Conceptualization, Methodology, Writing – Review and Editing, Supervision

TQA: Conceptualization, Methodology, Writing – Review and Editing, Supervision

LED: Conceptualization, Methodology, Writing – Review and Editing, Supervision

DNTT: Methodology, Formal analysis, Visualization, Writing – Review and Editing

KMY: Conceptualization, Methodology, Writing – Original Draft, Writing – Review and Editing, Visualization, Resources, Supervision, Funding Acquisition, Software

